# Detection of malaria parasites in dried human blood spots using mid-infrared spectroscopy and logistic regression analysis

**DOI:** 10.1101/19001206

**Authors:** Emmanuel P. Mwanga, Elihaika G. Minja, Emmanuel Mrimi, Mario González Jiménez, Johnson K. Swai, Said Abbasi, Halfan S. Ngowo, Doreen J. Siria, Salum Mapua, Caleb Stica, Marta F. Maia, Ally Olotu, Maggy T. Sikulu-Lord, Francesco Baldini, Heather M. Ferguson, Klaas Wynne, Prashanth Selvaraj, Simon A. Babayan, Fredros O. Okumu

## Abstract

**Background:** Epidemiological surveys of malaria currently rely on microscopy, polymerase chain reaction assays (PCR) or rapid diagnostic test kits for *Plasmodium* infections (RDTs). This study shows that mid-infrared (MIR) spectroscopy coupled with supervised machine learning could constitute an alternative method for rapid malaria screening, directly from dried human blood spots.

**Methods:** Filter papers containing dried blood spots (DBS) were obtained from a cross-sectional malaria survey in twelve wards in south-eastern Tanzania in 2018/19. The DBS were scanned using attenuated total reflection-Fourier transform infrared (ATR-FTIR) spectrometer to obtain high-resolution MIR spectra in the range, 4000 cm^-1^ to 500 cm^−1^. The spectra were cleaned to compensate for atmospheric water vapor and CO2 interference bands and used to train different classification algorithms to distinguish between malaria-positive and malaria-negative DBS papers based on PCR test results as reference. The analysis considered 296 individuals, including 123 PCR-confirmed malaria-positives and 173 negatives. Model training was done using 80% of the dataset, after which the best-fitting model was optimized by bootstrapping of 80/20 train/test stratified splits. The trained models were evaluated by predicting *Plasmodium falciparum* positivity in the 20% validation set of DBS.

**Results:** Logistic regression was the best-performing model. Considering PCR as reference, the models attained overall accuracies of 92% for predicting *P. falciparum* infections (specificity = 91.7%; sensitivity = 92.8%) and 85% for predicting mixed infections of *P. falciparum* and *P. ovale* (specificity = 85%, sensitivity = 85%) in the field-collected specimen.

**Conclusion:** These results demonstrate that mid-infrared spectroscopy coupled with supervised machine learning (MIR-ML) could be used to screen for malaria parasites in dried human blood spots. The approach could have potential for rapid and high-throughput screening of *Plasmodium* infections in both non-clinical settings (e.g. field surveys) and clinical settings (diagnosis to aid case management). However, full utility will require further advances in classification algorithms, field validation of this technology in other study sites and an in-depth evaluation of the biological basis of the observed test results. Training the models on larger datasets could also improve specificity and sensitivity of the technique. The MIR-ML spectroscopy system is robust, low-cost, and requires minimum maintenance.

## Background

Accurate diagnosis of malaria infection in both humans and mosquitoes is essential for understanding transmission patterns, estimating epidemiological burden and informing appropriate management of cases. The WHO’s Global Technical Strategy for Malaria 2016-2030 (GTS) recommends that surveillance strategies should be integrated as core interventions to provide better estimates of disease burden, improve resource allocation and accelerate progress towards elimination [1]. The malERA Refresh Consultative Panel on Tools for Malaria Elimination reiterated in 2017 that new diagnostic tools are as important as new drugs, vaccines and vector control tools [2].

Given health system challenges facing many low-income malaria endemic countries, there is particular interest in non-immunological point-of-care (POC) techniques that could be readily scaled up with minimum effort [2]. Equally vital is the need for better quantification and identification of asymptomatic pathogen carriers in situations of low transmission and sub-microscopic parasitaemia [2,3]. Low-cost mass screening tools are also vital for large cross-sectional investigations such as national malaria indicator surveys, and for monitoring progress of interventions such as insecticide-treated bed nets [4–6], indoor residual spraying [7], larviciding [8] and antimalarial medicines [9].

Malaria diagnosis from human samples currently relies on light microscopy with thin and thick blood smears [10], polymerase chain reaction assays [11–13] and a variety of antigen-detecting rapid diagnostic tests [14]. As countries approach elimination, there is greater need for accurate detection of malaria parasites in both symptomatic and asymptomatic individuals [15]. Microscopy is still commonly used in diagnosis to support case management [10]. With highly-trained and experienced microscopists, it can offer reliable results and enable identification and quantification of sexual and asexual stages of infections by different malaria parasite species (*Plasmodium falciparum, P. vivax, P. malariae* and *P. ovale*, being the most prevalent). However, microscopy-based diagnosis is labor-intensive and often requires more than one highly-trained person for confirmation of conflicting results [10]. Additionally, this technique has low sensitivity in many laboratories and misses most low-level parasitaemia cases [16].

PCR, on the other hand, is highly specific and sensitive for malaria parasite detection, but is not widely used for primary diagnosis in most malaria endemic places because it is expensive, requires highly skilled personnel, and is therefore impractical to implement in rural and remote facilities [17]. Furthermore, until the impact of treating submicroscopic parasitaemia on transmission is fully understood, WHO has recommended that PCR should not be part of routine malaria control or elimination programs [18]. It is however capable of identifying low-level parasitaemia that are otherwise undetectable by other methods, and distinguishing between individual parasite species. Recent developments in PCR applications have signaled the potential of non-invasive malaria diagnostic options such as those relying on DNA detection in saliva, urine, sweat and even fecal excreta [19–21]. Besides, real time PCR assays enable quantitative assessments and comparison of infection loads [15], but are expensive for most laboratories. A related technology is the loop-mediated isothermal amplification (LAMP), which is also increasingly used for diagnosis of multiple malaria species, and can be conducted at room temperatures without PCR instruments [22].

At present, malaria rapid diagnostic tests (mRDTs) are the best option for addressing the technical limitations of both light microscopy and DNA-based diagnosis. These tests target persistent specific antigens from malaria parasites [14,23,24] and are quick and deployable at large-scale. They also do not require highly-skilled labor, electricity or sophisticated storage needs. Unfortunately, mRDTs can be unreliable in low transmission settings, resulting in significant false negative and false positive results [25]. Both microscopy and mRDTs are recommended only when the number of malaria parasites exceed 100/µL, and are therefore not applicable for measuring low-level parasitaemia or identifying asymptomatic cases [16]. In one example, mass mRDT screening followed by treatment did not result in reduction of malaria incidence in the pre-elimination settings of Zanzibar, most likely due to poor reliability of the mRDTs [26]. However, a new rapid lateral flow technique that detects specific proteins of the infectious *Plasmodium* stages, i.e. gametocytes, in saliva is showing encouraging results in detecting sub-microscopic parasites in children and adults [27].

Recent studies have shown that non-molecular techniques such as near-infrared spectroscopy (12,500 cm^−1^ to 400 cm^−1^ frequencies) and mid-infrared spectroscopy (4000 cm^−1^ to 400 cm^−1^ frequencies), combined with advanced data analysis, could provide cheaper, quicker, reagent-free and potentially simpler options for surveys of mosquitoes and mosquito-borne infections. Examples include detection of endosymbionts such as *Wolbachia* bacteria, and pathogens such as *Plasmodium* and Zika virus in mosquitoes [28–31]. Such approaches have also been used for estimating age of disease-transmitting mosquitoes [32–38], distinguishing between vector species [32,38,39] and assessing their blood feeding histories [40], all of which directly influence malaria transmission.

Khoshmanesh *et al*., [41] used mid-infrared (MIR) spectroscopy combined with partial least-squares (PLS) regression to detect early ring stages of laboratory-cultured *P. falciparum* with detection limits less than 1 parasite/µL. To improve this approach, Roy *et al*., [42] used *Plasmodium* cultures to spike whole blood obtained from six uninfected volunteers, then aliquoted these mixtures multiple times to obtain 132 specimens containing different quantities of *P. falciparum* parasites, glucose and urea. Based on PLS regressions analysis of MIR spectra from these aliquots, they correctly identified 98% of specimens with parasitaemia densities above 0.5% (∼25,000 parasites/µL). Sensitivity was however only 70%, possibly because the model included only a small number of negative samples. Though limited to laboratory cultures and small number of samples with low genetic variability, these studies were the first to demonstrate direct potential of MIR for malaria parasite detection.

This current study has extended the approach used by Khoshmanesh *et al*., [41] and Roy *et al*., [42], to provide the first demonstration of MIR spectroscopy coupled with supervised machine learning (MIR-ML) to diagnose malaria in dried human blood spots (DBS) obtained from field surveys of naturally infected individuals in a malaria-endemic community in Tanzania.

## Methods

### Study Area

Samples for this analysis were obtained from a cross-sectional malaria parasite prevalence survey conducted in 12 administrative wards in Ulanga and Kilombero districts in south-eastern Tanzania (Fig. 1). Average altitude of the area is 270m above sea level, annual rainfall is 1200-1800 mm and annual temperatures range between 20°C and 32°C. Malaria prevalence at the time of the survey varied from <1% in the northernmost urban wards of Ifakara town and Mlabani, to >40% in southern wards of Igota and Igumbiro (Swai et al., unpublished). *P. falciparum* accounts for more than 90% of known malaria parasites in the area.

**Fig. 1:**
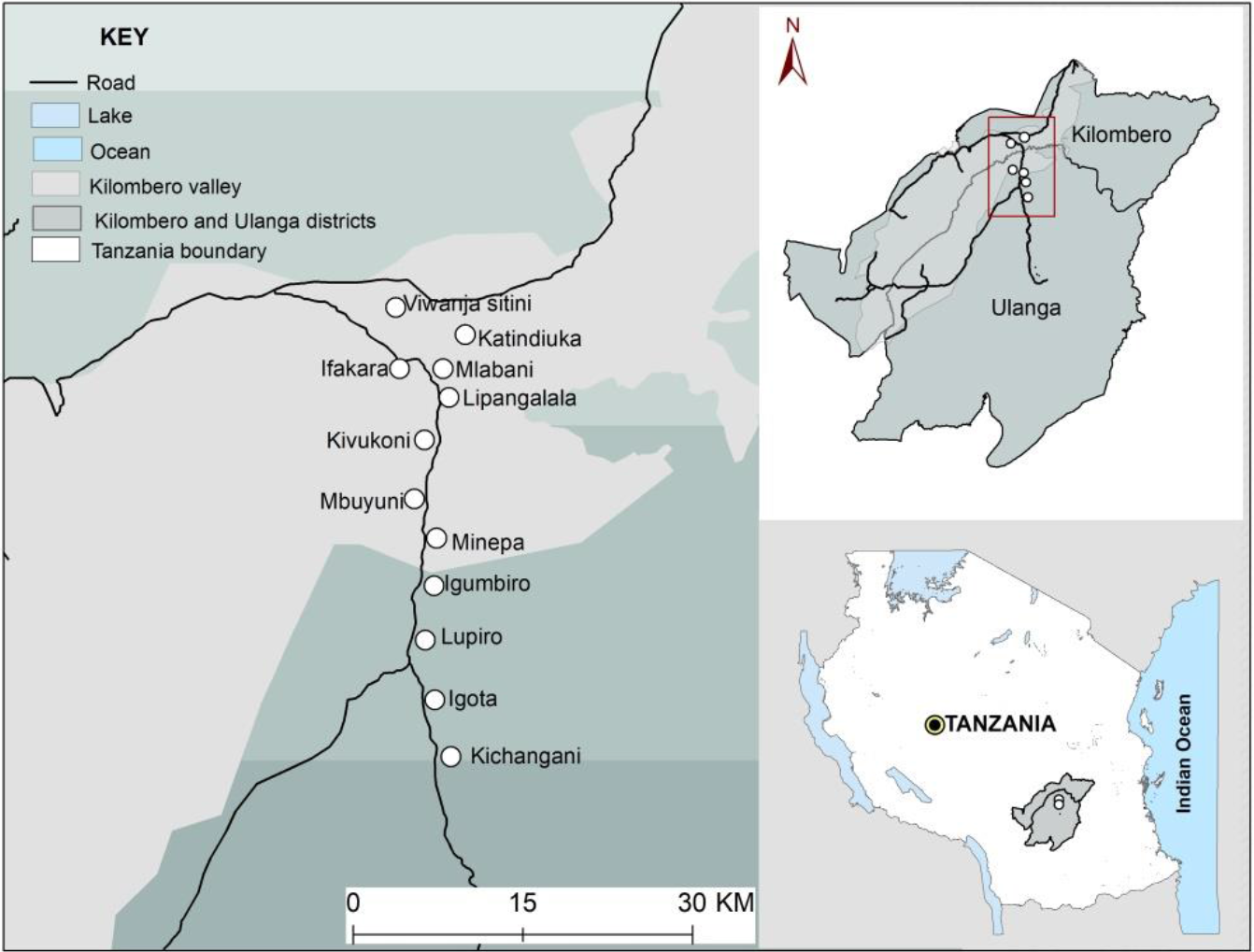
Map showing study villages in Kilombero and Ulanga districts, south-eastern Tanzania (courtesy of Alex J. Limwagu).

Transmission intensities also vary and were last estimated to be ∼16 infectious bites/person/year in the Ulanga district wards [43], and <1 infectious bites/person/year in the Kilombero district wards [44]. The dominant vector species is *Anopheles funestus* followed by *An. arabiensis* [43]. Long-lasting insecticidal treated bed nets are the most common prevention tool used in the study with coverage of > 80% [4].

### Malaria parasite surveys using mRDTs

The parasite survey included 1,486 households and 3,292 individuals across the 12 study wards (Fig. 1), and was conducted between September 2018 and February 2019. Blood samples were taken from finger pricks of household members of all age groups and both sexes (Swai *et al*., unpublished), and then tested for malaria parasites using mRDTs and polymerase chain reaction (PCR).

The mRDTs (SD BIOLINE malaria Ag P.f/Pan 05FK60) were used to screen all individuals and results interpreted following manufacturer’s instructions [45]. This mRDT kit detects *P. falciparum* and other *Plasmodium* species in whole blood samples [45] by differential detection of circulating histidine-rich antigen II (HRP-II) of *P. falciparum*, and common *Plasmodium* lactate dehydrogenase (pLDH) of *Plasmodium* species [14,23,24]. All malaria positive specimens were verified by light microscopy, and if found positive, the individual was immediately treated by a study clinician according to Tanzania national treatment guidelines for malaria [46].

In addition to microscopy and mRDTs, dry blood spot (DBS) specimens were collected from all mRDT-tested patients regardless of test results. Three to four drops of blood were placed directly from finger prick onto Whatman 903™ card (GE Healthcare Bio-Sciences Corp.) [47]. The DBS specimens were dried in open air for 15 minutes, and stored in zip-locked plastic bags, with desiccants. The DBS were stored at −20°C for 2-5 months, after which further tests were done, all at once, as described below.

### Mid-infrared scanning of dry blood spots

The DBS collected from individuals for which there had been a positive malaria test results (n = 147), and a separate random selection of specimens from individuals with negative results (n = 149) were scanned using a Bruker ALPHA Fourier-Transform Infrared (FTIR) spectrometer equipped with Platinum ATR and a Platinum diamond sampling module [48]. The individual DBS cards were placed between the anvil and crystal plate of the spectrometer, each time ensuring that the MIR beam directly strikes the middle of the blood spots (Fig. 2a).

**Fig. 2:**
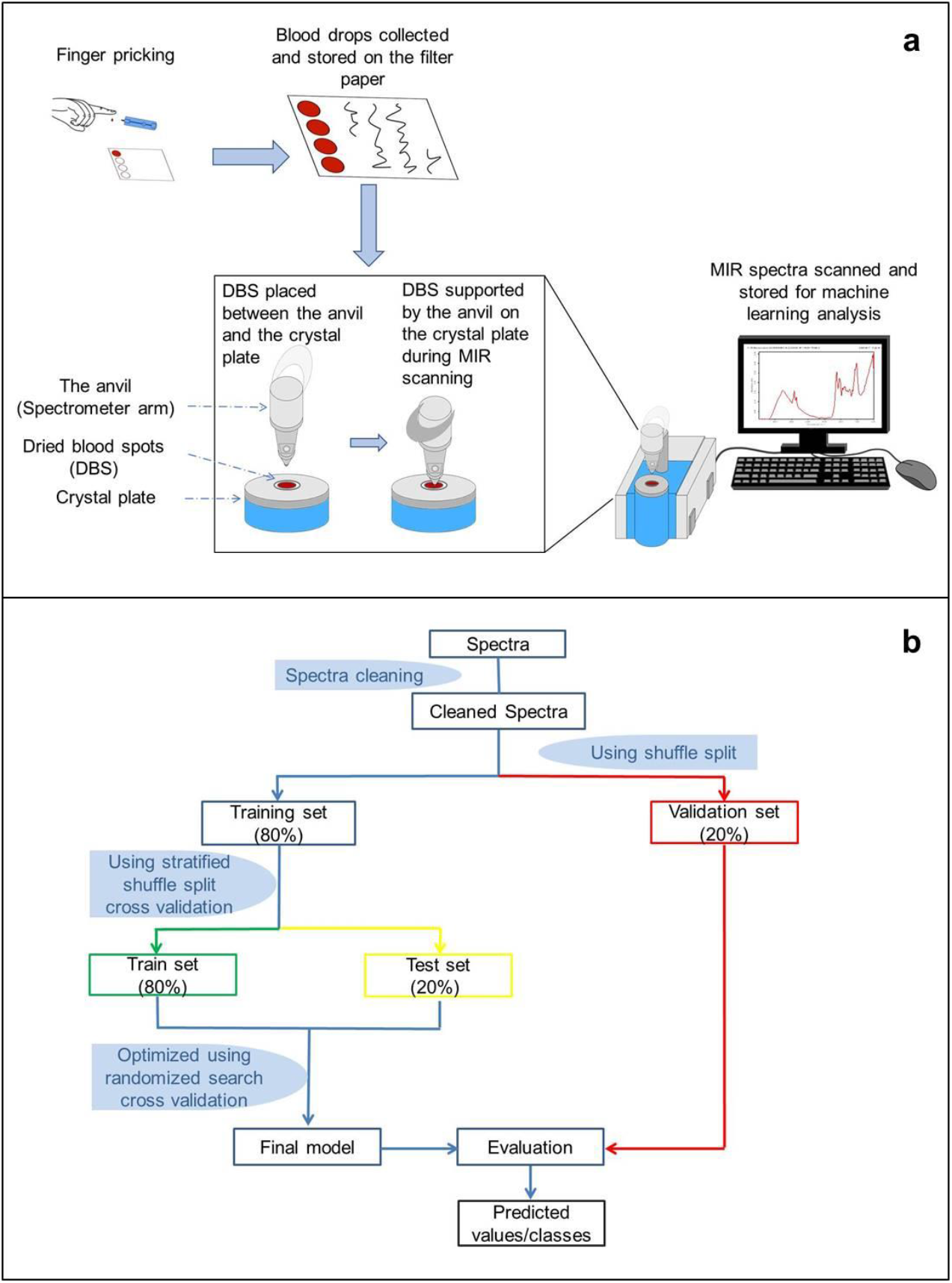
Schematic Illustration specific processes of: (a) collection of blood specimens and preparation of dried blood spots (DBS) on filter papers, scanning on mid-infrared spectrometer, and recording sample spectra, (b) data splitting, model training, cross-validation and evaluation of performance of final model (supervised machine learning process).

The MIR spectra were captured from 4,000 to 500 cm^−1^ wavenumbers, with spectral resolution set at 5 cm^−1^. Each individual DBS sample was scanned 32 times in 30 seconds, and averaged to obtain a single representative spectrum. The anvil and crystal were cleaned after each specimen scan, using velvex tissue soaked in 70% ethanol and dried using a clean dry tissue. The spectrometer has an internal validation unit (IVU) with reference standards, and was programmed to conduct automated instrument tests for operational and performance qualification. However, additional performance and quality checks were done by conducting frequent background scans before first sample scan and after every five specimen scans. Additional details of the MIR spectrometer and the scanning process have been described elsewhere [38,40]. All DBS samples were scanned at the same time.

### Nested PCR assays to confirm malaria infections in the dry blood spots

DNA was extracted from the DBS papers and the presence of any of the five malaria parasites (*P. falciparum, P. ovale, P. vivax, P. malariae* and *P. knowlesi*) examined using nested PCR targeting 18S ssRNA sequences, according to the Snounou *et al*., protocol with slight modification [49]. The first PCR reaction used 10µl of DNA, amplified with rPLU1 and rPLU5 primers targeting fragment lengths between 1600bp and 1700bp. The second PCR stage used 2µl of the first PCR product, plus species-specific primer pairs (rFAL1-rFAL2, rOVA1-rPLU2, rVIV1-rVIV2, rMAL1-rMAL2, and Pmk8-Pmkr9) in five different reactions. Amplification for all reactions was done in 20µl PCR master mix. The PCR products were analyzed after electrophoresis in agarose gel and DNA bands visualized using Kodak Logic 100 imaging system.

### Data analysis

The OPUS software [50] was used to clean and compensate all spectra for water vapor absorption bands and carbon dioxide (CO2) interference bands as described in Mwanga *et al*., [40] and Gonzalez-Jimenez *et al*., [38]. A final dataset of 296 samples, including 123 PCR-confirmed *Plasmodium* positive and 173 *Plasmodium* negative individuals was considered for further analysis in the Python programming language version 3.7 with the *scikit-learn* library [51]. All wavenumbers from the spectra were included in the analysis. Supervised machine learning algorithms were trained to map spectra to known PCR test results, by implementing binary classification strategy, and validated on a separate set of spectra not used for training (Fig. 2b).

Primary assessment was done against PCR as reference because of its high sensitivity and specificity compared to mRDTs and microscopy [17,52]. At this proof-of-concept stage, it was however not possible to balance the specimens by other variables such as anemia, sex, age, period of storage or prevailing parasitaemia prevalence in the different villages.

The PCR analysis had identified mostly infections of *P. falciparum* occurring singly, though there were also a few cases of mixed infections consisting of *P. falciparum*, together with *P. ovale*. The MIR-based predictions were therefore done first by excluding the mixed infections, and second, without excluding them.

### Classifying DBS samples as positive or negative for *P. falciparum*, based **on** PCR test results as reference

All samples with mixed *Plasmodium* species infections (N = 9) were excluded, so that the remaining dataset included only *P. falciparum* positives and a reduced number of negative samples (N = 260; N _positives_ = 114, and N _negatives_ = 146). As previously described [40], the dataset was first partitioned into training set (80%; N = 208) and validation datasets (20%; N = 52). The validation set was preserved for evaluating prediction accuracies of the final model.

The training dataset was subjected to multiple rounds of randomly stratified splits into training sets (80%) and test sets (20%), to achieve rigorous classification of the different malaria-positive and malaria-negative samples. To provide a mix of data representation methods and allow for both parametric and non-parametric assessment of the data, baseline performances of seven different classification algorithms were tested. These included: K-nearest neighbors classifier (KNN), logistic regression (LR), support vector machine classifier (SVM), naïve Bayes (NB), random forest classifier (RF), XGBoost classifier (XGB), and multilayer perceptron (MLP) [51]. All code was adapted from https://github.com/SimonAB/Gonzalez-Jimenez_MIRS.

The algorithms were trained on spectral features representing malaria-positive and malaria-negative DBS samples (Fig. 3). The best performing classification algorithm was optimized by fitting 70 bootstrapped models, for which the outputs were aggregated to obtain an ensemble model with highest accuracy and lowest variance. Finally, best performing classifiers were bagged and used to compare the predicted labels against true labels (*Plasmodium* positive or *Plasmodium* negative) using the naïve validation set separated at the beginning.

**Fig. 3.**
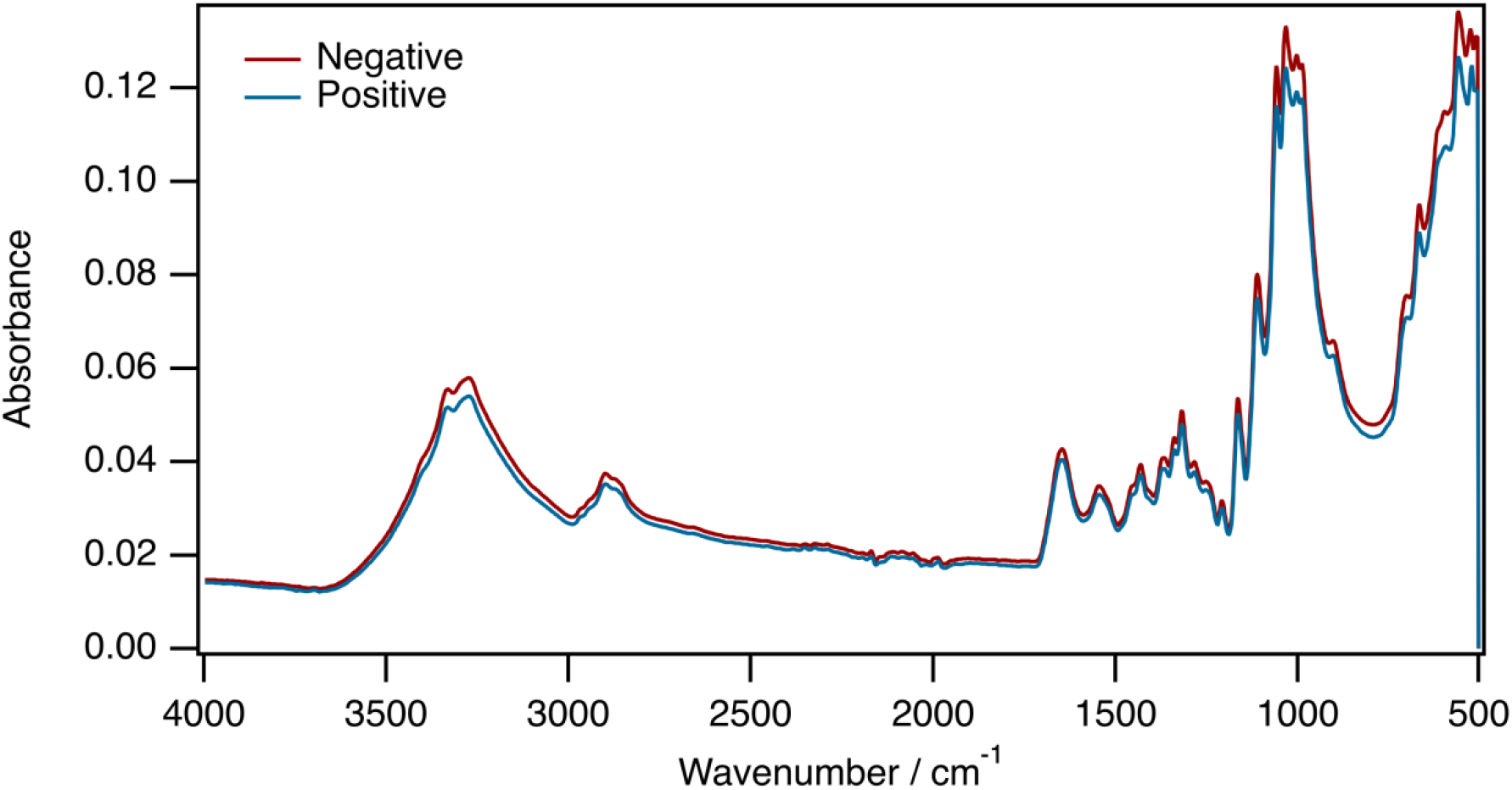
Averaged mid-infrared spectra obtained from dried blood spots confirmed by PCR as *Plasmodium* positive or *Plasmodium* negative. Assignations of biochemical properties of different wavelengths are shown in Table 1.

The full analysis was repeated, this time including the mixed infections, which had been removed from the initial model training. This way, the MIR-ML approach was evaluated for predicting infection with any of the two *Plasmodium* species (*P. falciparum* and *P. ovale*) occurring singly or together in a DBS.

Lastly, based on the previous spectroscopic analysis of body fluids [53,54], and the earlier work by Roy *et al*. [42], Kozicki *et al*., [55], Khoshmanesh *et al*., [41], we examined putative bands possibly responsible for the spectral differences between *Plasmodium*-infected and *Plasmodium*-free specimen. The sensitivity of the candidate MIR bands was assessed for lipids, proteins, carbohydrates and nucleic acids. We also extracted prediction coefficients of the best model and plotted its most dominant features (wavenumbers).

## Results

### Spectral characteristics of dried blood spots with and without *Plasmodium*

Fig. 3 shows two averaged spectra obtained from dried blood spots confirmed by PCR as *Plasmodium* positive or *Plasmodium* negative, and the features (or wavenumbers) relied upon to predict the infection status. The specific biochemical properties associated with major peaks in the spectra are shown in Table 1. The detectable vibrational modes show a complex of potentially distinctive biochemical characteristics associated with lipids, proteins, amides, sugars and the heme group (Table 1).

**Table 1:** Biochemical properties associated with peaks in the mid infrared spectra obtained from dried blood spots in Fig. 3.

| Wavenumber (cm<sup>-1</sup>) | Vibrational mode | Component identification |
| --- | --- | --- |
| 3600 – 3000 | N-H stretching | Amides (proteins, hemoglobin), urea |
|  | O-H stretching | Alcohols carbohydrates, cellulose |
| 3332 | O-H stretching | Alcohol cellulose |
| 3293 | N-H stretching | Amide (proteins, hemoglobin) |
| 3272 | O-H stretching | Cellulose |
| 3000 - 2800 | C-H stretching | Lipids, amino acids, carbohydrates |
| 2894 | C-H stretching | Cellulose, carbohydrates |
| 1700 – 1600 | C=O stretching | Amides (proteins, hemoglobin), urea |
| 1540 | N–H bending<br>coupled to C–N<br>stretching | Amides (proteins, hemoglobin) |
| 1457 | CH<sub>3</sub> bending | Amino acids, lipids |
| 1400 – 1310 | C-H stretching | Lipids, amino acids, carbohydrates |
| 1307 | C-N stretching | Amides (proteins, hemoglobin) |
| 1165 – 1110 | C-O-C stretching | Ethers (cellulose, carbohydrates) |
| 1070 - 950 | C-O stretching | Alcohols (cellulose, carbohydrates, amino-acids) |
|  | =C-H bending | Heme group; hemoglobin |
| 730 | C-H bending | Lipids, amino acids, carbohydrates |

### Classifying DBS samples as positive or negative for *P. falciparum*, based on PCR as reference

In the first analysis, only *P. falciparum* infected individuals were considered in the category of malaria positive DBS cards, and any mixed infections were excluded. Logistic regression (LR) outperformed the other six candidate classifiers in accuracy and precision (Fig. 4a). The best LR models predicted *P. falciparum-*infected and non-infected individuals with overall accuracy up to 90% before optimization (Fig. 4a) and 91.5% after optimization (Fig. 4b & 5a). When challenged with the validation dataset, the optimized model achieved an overall accuracy of 92.3%, correctly identifying 92% of PCR-negative individuals and 93% of PCR-positive individuals (Fig. 5b).

**Fig. 4:**
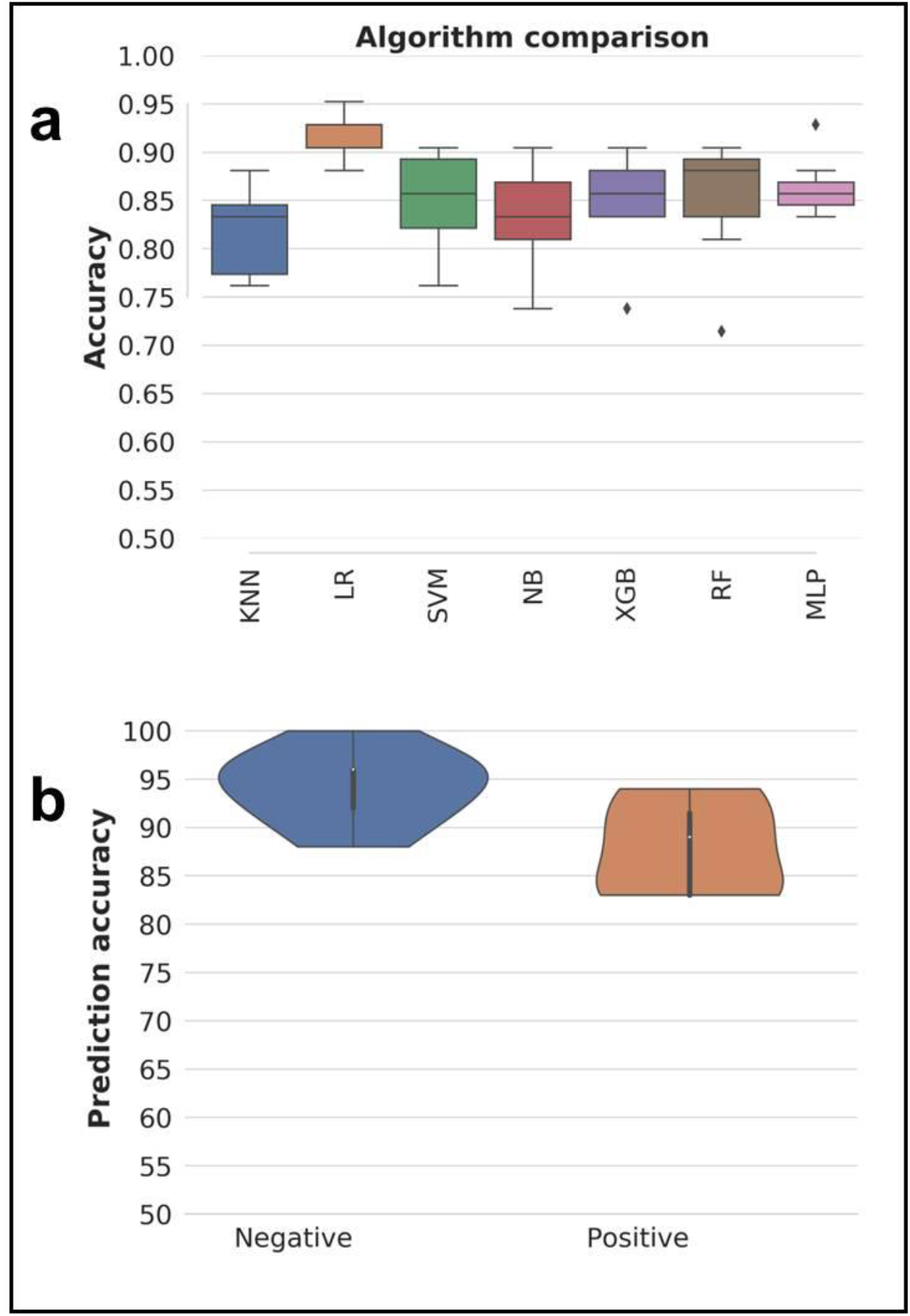
**(a)** Prediction accuracies and precision for different classification models, based on PCR test results as reference. Models compared included k-nearest neighbors (KNN), logistic regression (LR), support vector machines (SVM), naïve Bayes (NB), XGBoost (XGB), random forest (RF), Multilayer perceptron (MLP). Logistic regression (LR) was the best performing model; (**b**) distribution of per class accuracies obtained by final LR classifiers and standard deviation from 70 bootstrapped models in predicting PCR test results from MIR spectral data.

**Fig. 5:**
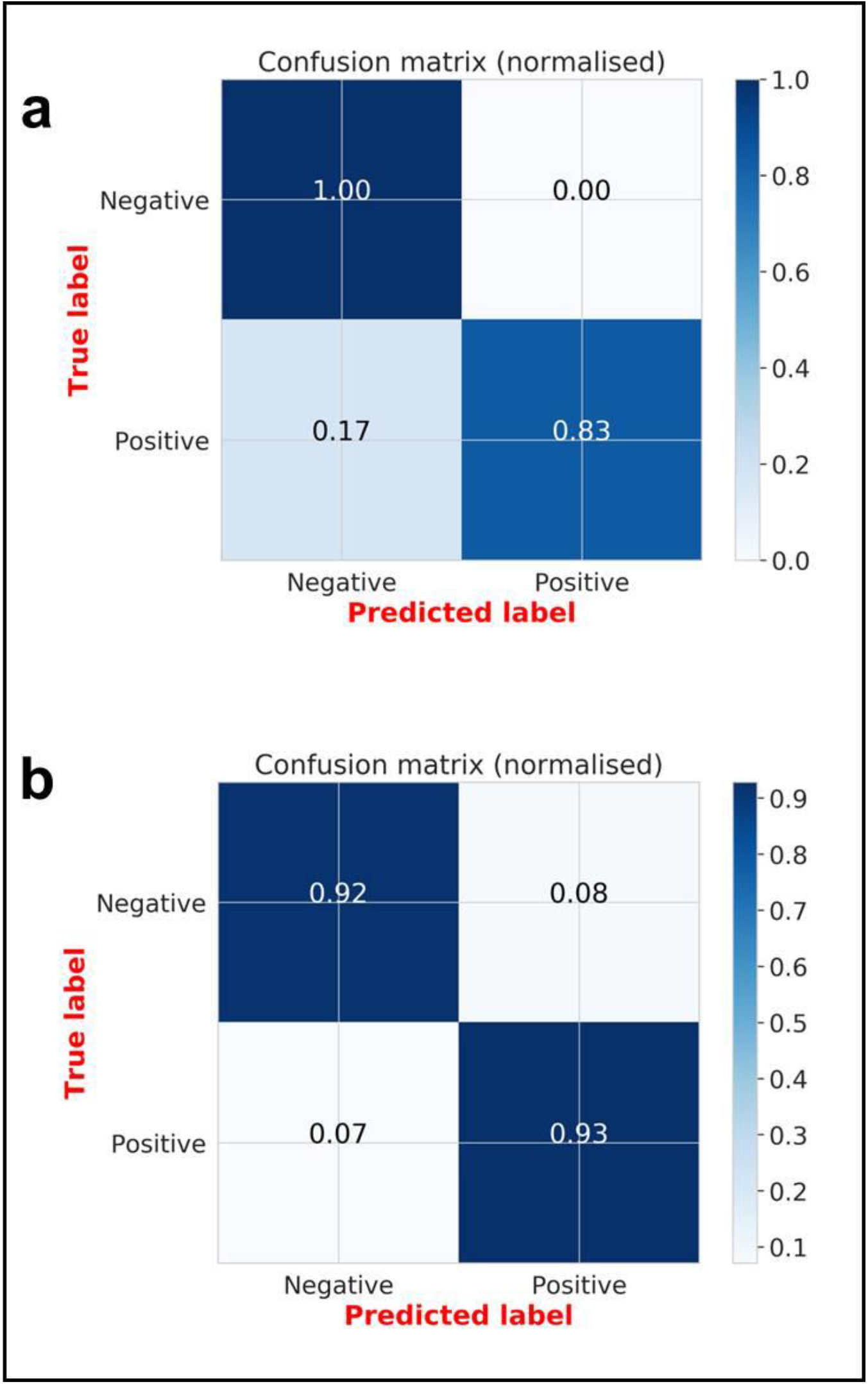
**(a)** averaged proportions of correct predictions of PCR-confirmed *Plasmodium falciparum* infected individuals achieved during the training of the models; **(b)** averaged proportions of correct predictions of *P. falciparum* infected individuals achieved when the final model is challenged the previously unseen validation spectra.

The validity of MIR-ML, is shown in Table 2. The approach was 92.8% sensitive and 91.7% specific. For comparison, the validity of the mRDTs used in the survey, when compared to PCR is also shown. The mRDTs were 97.6% sensitive and 84.4% specific for detection of *Plasmodium*-infected individuals in this study area.

**Table 2:** Performance of mid-infrared spectroscopy coupled with logistic regression, and the mRDT (i.e. SD BIOLINE malaria Ag P.f/Pan 05FK60), both compared to PCR for identifying *Plasmodium falciparum* infected individuals from the validation set.

|  |  | PCR |  | Total | % Sensitivity | % Specificity | % Positive Predictive Value | % Negative Predictive Value |
| --- | --- | --- | --- | --- | --- | --- | --- | --- |
|  |  | Positive | Negative |  |  |  |  |  |
| Mid-Infrared Spectroscopy & Machine Learning [MIR-ML] | Positive | 26 | 2 | 28 | 92.8 | 91.7 | 92.8 | 91.7 |
|  | Negative | 2 | 22 | 24 |  |  |  |  |
|  |  | **28** | **24** | **52** |  |  |  |  |
|  |  | **Positive** | **Negative** | **Total** | **% Sensitivity** | **% Specificity** | **% Positive Predictive Value** | **% Negative Predictive Value** |
| SD BIOLINE malaria Ag P.f/Pan 05FK60 mRDT | Positive | 120 | 27 | 147 | 97.6 | 84.4 | 86.0 | 97.2 |
|  | Negative | 3 | 146 | 149 |  |  |  |  |
|  |  | **123** | **173** | **296** |  |  |  |  |

### Selection of most dominant spectral features used for distinguishing between *P. falciparum* positive or negative DBS

The LR coefficients showed the relative importance of wavenumbers, enabling selection of those most responsible in the decision function (prediction of malaria positive and malaria negative DBS). Fig. 6 therefore shows the wavenumbers with the strongest likelihood in predicting the infection status of the individuals. All the top twenty wavenumbers (features) were in the fingerprint region of the MIR spectra (1730 cm^−1^ −883 cm^−1^) where lipids, amino acids and proteins is mostly detected (Table 1)

**Fig. 6:**
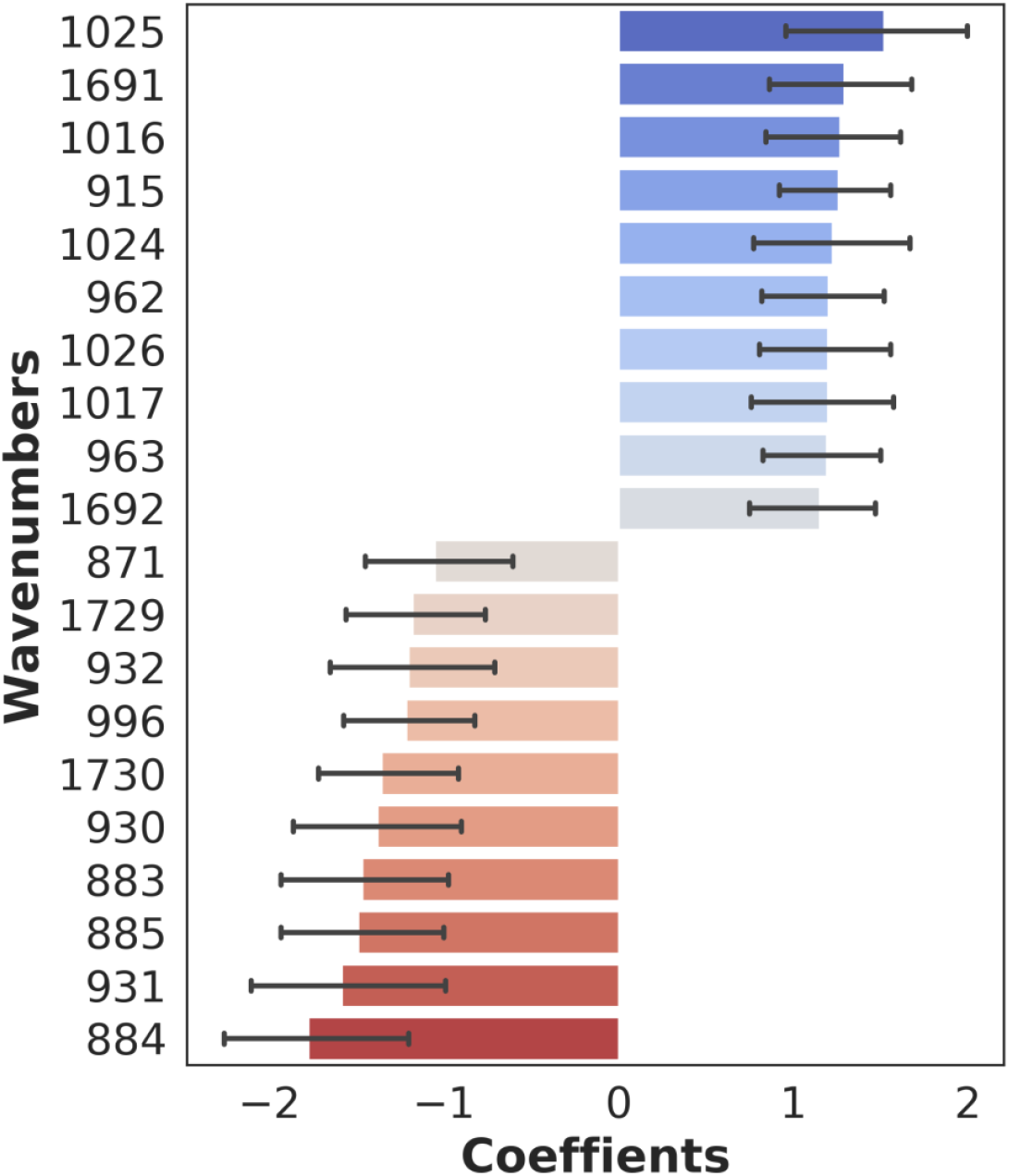
Plots of twenty most dominant spectral features (wavenumbers) influencing model prediction of *Plasmodium*-infection status of the DBS specimens. The positive coefficients are those most predictive of *Plasmodium* positive specimens while the negative coefficients are those most predictive of *Plasmodium*-negative specimen. All the top twenty features are found in the fingerprint region (1730 cm^−1^ - 883 cm^−1^)

### Predicting PCR test results from MIR spectra without excluding mixed infections

In the second analysis, all infected individuals were considered regardless of *Plasmodium* species present. Once again, LR was the best model, achieving up to 93% accuracy and 95% after bootstrapping. The final model achieved 87% overall accuracy for the validation set, correctly identifying 91% of PCR-negative individuals and 82% of PCR-positive individuals.

## Discussion

This study is an initial demonstration using field-collected blood specimens that MIR spectroscopy coupled with logistic regression analysis, can be harnessed for detection of infectious parasitic diseases, in this case malaria. Malaria cases were identified and analyzed by both MIR spectroscopy and PCR, then the results were compared. As a proof of principle, it considered 296 individual samples, to demonstrate potential application of MIR spectroscopy for malaria detection. The main finding was that spectral signatures collected from dried human blood spots can be relied upon to identify malaria-infected and non-infected specimens.

Validity of this approach was verified by PCR tests, and corroborates the earlier evidence by Khoshmanesh *et al*. [41] and Roy *et al*. [42], who first demonstrated applications MIR spectroscopy to detect *Plasmodium*. Other related approaches include the use of surface-enhanced Raman spectroscopy (SERS), requiring silver nanoparticles to be added to lysed blood, and magneto-optic scanning to detect hemozoin, a waste product of *Plasmodium* infection of red cells pigment, haemoglobin [56,57]. Both of these approaches have been tested *in vitro*, though Newman *et al*., also evaluated the magneto-optic systems in a small preclinical trial with 13 participants [58].

An important advancement in the work presented here is that it has demonstrated the first direct application of MIR spectroscopy on field-collected dried blood specimens on filter papers. This approach makes the technique readily applicable without any additional reagents or pre-processing of samples. The 296 samples analyzed came from multiple age groups of both male and females in villages with varying malaria prevalence rates, thereby providing considerable variability between individual infections. This further emphasizes the potential of this technology in real-world settings for epidemiological screening, or diagnostics for patient management. When the final optimized LR model was applied to new blood samples that had not been exposed to the classifier, they correctly identified 92% of the malaria-free individuals and 93% of malaria-infected individuals compared to PCR. The need for no additional sample preparation or reagents is particularly important for field practicality.

This study adds to the growing evidence showing the potential role of infrared spectroscopy and chemometrics in surveillance of mosquito-borne diseases. In a recent study conducted using near-infrared rather than mid-infrared spectra, it was possible to detect infectious *P. falciparum* sporozoites in laboratory-reared and laboratory-infected *Anopheles* mosquitoes with up to 90% accuracy [31]. Earlier studies had also demonstrated detection of various pathogens such as *Wolbachia, Plasmodium* and Zika virus in different mosquito species [28–31]. Many of these earlier approaches relied on spectroscopy at near-infrared (NIR) frequencies (12,500 cm^−1^ to 400 cm^−1^) where the absorption intensity is due to overtone and combination bands, which are relatively weak. Spectroscopy at mid-infrared (MIR) frequencies (4,000 cm^−1^ to 400 cm^−1^) captures the fundamental vibrational modes of biological samples, which are stronger and more information rich [38], thereby offering stronger signals than NIR, which reflects secondary modes. Advances in data analysis and machine learning techniques now make the large spectral datasets amenable to processing.

The distinctive features of the MIR spectra are likely related to the biochemical changes in red blood cells following malaria infection [41,42]. These differences may also result from *Plasmodium*-specific proteins present in infected human blood [59], or simply due to pathological manifestations of malaria such as anemia, iron deficiency or other inflammatory responses. Based on previous analyses of body fluids [53,54,60,61], and earlier work by Roy *et al*., [42], Kozicki *et al*., [55], Khoshmanesh *et al*., [41], the spectral bands putatively responsible for the differences between *Plasmodium*-infected and *Plasmodium*-free specimens were examined (Table 1). Further analysis of the dominant spectral wavelengths showed that they were mostly in the fingerprint region associated with amino acids, carbohydrates, lipids and proteins (Table 1 and Fig.6). More detailed analysis beyond this initial proof of principle is still required to examine how variations in these specific bands influence the diagnostic capacity of this technology.

Bands in the C−H stretching regions are usually attributed to lipids synthesized during development of *P. falciparum* and can change the make-up of infected erythrocytes and become detectable in the spectra. On the other hand, the region between 1,250 and 800 cm^−1^ is sensitive to nucleic acid vibrations associated with proliferation of *Plasmodium*-specific ribosomes during parasite development and cell invasion [42]. We observed several peaks in this region characteristic of sugars and peptides, consistent with parasite DNA (Table 1 & Fig. 6). In previous assessments, it was also shown that reduced absorbance in the carbohydrate regions at 1,144 cm^−1^, 1,101 cm^−1^ and 1,085 cm^−1^ are likely associated with lower glucose content in infected red blood cells, since these parasites metabolize glucose faster than normal cells [41,42]. Perhaps the most obvious are the different heme vibration regions (Table 1), which show higher hemoglobin levels in non-infected than infected samples because *Plasmodium* catabolizes the complex hemoglobin protein into constituent lipids and bilirubin. By tracking these key characteristics, it is possible to predict the infection status of the dried whole blood specimen.

This study should be considered purely as an initial proof-of-principle study, and more studies are required before the technology is field-deployable or effective. Filter papers containing dried human blood spots were scanned and the resulting spectra used to train algorithms to predict outcomes of nested PCR [40]. Comparison was made with results of tests considering only *P. falciparum* and also with results of tests considering any species of *Plasmodium*. In both cases, the technique achieved high accuracy in distinguishing between infected and uninfected specimen. While this finding indicates that the technique could be applicable for diagnosis of different malaria parasites, it also highlights the need to improve the approach so as to distinguish between different parasite species.

The field survey from which the DBS specimens were collected also had mRDT data. Relative to PCR as reference, the mRDT results achieved slightly higher sensitivity (97.6% vs 92.8%) but lower specificity (84.4% vs. 91.7%) than the mid infrared spectroscopy approach (Table 2). In previous studies, the same mRDTs achieved sensitivity of 99.7% for *P. falciparum*, and 95.5% for non *P. falciparum* and a specificity of 99.5% for both [45,62].

Significant improvements are still necessary before this technique can be deployed for actual screening and diagnosis. One avenue of improvement is the availability of more field data to validate this approach as a complementary tool for screening malaria, and potentially other blood-borne infections. Also, this current study did not include any quantification of infection intensities, which will be necessary to examine validity in areas of different transmission intensities, as well as potential role of this technology in malaria elimination settings. Such future studies should also include: a) greater analysis on biological basis of the observed signals and how this may be influenced by the natural history of malaria infections in humans, or its manifestations such as anemia, b) determination of whether the method can distinguish between the different parasite stages, such as the asexual stages versus the sexual stages (*i*.*e*. gametocytes), c) detailed examination and characterization of cases incorrectly identified by this new approach, and d) assessment of parasite detection thresholds, and factors that may influence it if mid-Infrared spectroscopy and machine learning are used.

It takes 30 seconds to scan a single blood spot under MIR spectrometer, and the approach is considerably lower cost than PCR platforms. The MIR equipment presently costs ∼29,000 USD as an initial outlay, but there are no extended costs for reagents except for occasional replacement of the desiccants. It takes less than a minute to clean the crystal and anvil, position DBS on the crystal and collect the MIR spectra, thus any experienced staff member can scan more than 250 specimens per day. Comparatively, average PCR systems cost between 3000USD and 10,000 USD, require reagents repeatedly. The cost of processing a DBS sample can be 2-4 USD per unit and regularly takes up to two days get back full results in batches typically not exceeding 100 specimens. Assuming a modest analysis of just 5000 samples per year, it would take ∼2 years to recover full costs of switching from PCR to MIR-ML based systems and break even. The onward costs of servicing are also low as the MIR equipment is robust and requires no reagents other than desiccants. Compared to PCR, use of MIR-ML could therefore potentially be developed into a cost-effective, quick and scalable approach. Besides, the same spectra, once collected, can be analyzed for multiple characteristics, potentially making this approach a one-stop system for assessing multiple disease indicators with different specimen types such as DBS papers, blood slides, mosquitoes and fluids. It is however not expected that the technology, in its current form, can replace current best practices for malaria surveillance or diagnostics, without further field validation.

One limitation of this current study is that the number of samples used was low, totaling only 296 (123 *Plasmodium* positive and 173 *Plasmodium* negative). It is expected that the quality of the predictions will improve as more data are available to train the models, especially if there is variation in localities from where the specimen originates, parasite densities, geographical locations and demographics of infected individuals. A related limitation was that it was not possible to balance the *Plasmodium*-positive and *Plasmodium*-negative specimens by other variables such as anemia, sex, age, period of storage or parasitaemia prevalence in the different villages, all of which may influence malaria risk and the predictive values of test methods. Future studies should therefore test whether these factors can significantly influence outcomes of the prediction models.

## Conclusions

These results demonstrate that mid-infrared spectroscopy coupled with supervised machine learning could be used to screen for malaria parasites in dried human blood spots. This approach has the potential for rapid and high-throughput screening of *Plasmodium* infections in non-clinical field surveys, and possibly for diagnosis in clinical settings. However, the full utility will require further advances in the algorithms and field validation in other study sites. Training the models on larger datasets is expected to improve specificity and sensitivity of the technique. The MIR spectroscopy system is robust, lower-cost compared to PCRs, requires minimum maintenance, and is reagent-free except for occasional replacement of desiccants.

## Ethics approval

Ethical approval for the study was obtained from Ifakara Health Institute’s Institutional Review Board (Ref. IHI/IRB/No: 007 - 2018), and from the Medical Research Coordinating Committee (MRCC) at the National Institutes of Medical Research (NIMR), Ref: NIMR/HQ/R.8a/Vol.IX/2895.

## Consent for publication

Permission to publish this work was also obtained from the National Institutes of Medical Research (NIMR), REF: NIMR/HQ/P.12 VOL XXVIII/14.

## Data Availability

All data for this study will be available upon request

https://github.com/MwangaEP/Mannu-ML-projects/tree/master/My%20projects/DBS%20work

## Availability of data and material

All data for this study will be available upon request. The analysis script was adapted from https://github.com/SimonAB/Gonzalez-Jimenez_MIRS; and has been changed to accommodate this specific analysis and deposited in a public repository. It is available at MwangaEP/Manuu-ML-Projects.

## Authors’ contribution

EPM, EM, EGM, HSN, SAM, DJS, JKS and FOO designed the study. EPM, EM, JKS and EGM performed and supervised experiments and collection of dried blood spots samples. EPM and MGJ analyzed the data. EPM and CS designed the schematics. SA and EM supported with PCR analysis. EPM, EM, EGM and FOO wrote and revised the manuscript. HMF, MFM, FB, SAB, KW, AO, MTS, and PS reviewed and revised the manuscript. SAB trained EPM the Machine learning analysis. The manuscript has been approved by all authors prior to submission. All authors read and approved the final manuscript.

## Acknowledgment

The authors wish to thank all field technicians and senior health personnel who supported malaria diagnosis and DBS collection. The authors thank Ifakara laboratory team for performing PCR analysis.

